# SARS-CoV-2 evolution and immune escape in immunocompromised patients treated with exogenous antibodies

**DOI:** 10.1101/2022.04.12.22273675

**Authors:** Erin M. Scherer, Ahmed Babiker, Max W. Adelman, Brent Allman, Autum Key, Jennifer M. Kleinhenz, Rose M. Langsjoen, Phuong-Vi Nguyen, Ivy Onyechi, Jacob D. Sherman, Trevor W. Simon, Hannah Soloff, Jessica Tarabay, Jay Varkey, Andrew S. Webster, Daniela Weiskopf, Daniel B. Weissman, Yongxian Xu, Jesse J. Waggoner, Katia Koelle, Nadine Rouphael, Stephanie M. Pouch, Anne Piantadosi

## Abstract

**Background:** SARS-CoV-2 mutations conferring escape from neutralizing antibodies can arise in immunocompromised patients with prolonged infection, but the conditions that facilitate immune escape are still not fully understood.

**Methods:** We characterized endogenous immune responses, within-host SARS-CoV-2 evolution, and autologous neutralization of the viral variants that arose in five immunocompromised patients with prolonged infection and B cell deficiencies.

**Results:** In two patients treated with the monoclonal antibody bamlanivimab, viral resistance to autologous serum arose early and persisted for several months, accompanied by ongoing evolution in the spike protein. These patients exhibited deficiencies in both T and B cell arms, and one patient succumbed to disease. In contrast, we did not observe spike mutations in immunologically important regions in patients who did not receive exogenous antibodies or who received convalescent plasma and had intact T cell responses to SARS-CoV-2.

**Conclusions:** Our results underscore the potential importance of multiple factors – the absence of an effective endogenous immune response, persistent virus replication, and selective pressure such as single-agent bamlanivimab – in promoting the emergence of SARS-CoV-2 mutations associated with immune evasion. These findings highlight the need for larger clinical studies in immunocompromised populations to better understand the ramifications of different therapies. Our results also confirm that patients with B cell deficiencies can elicit effector T cells and may suggest an important role for T cells in controlling infection, which is relevant to vaccines and therapeutics.

## Introduction

Immunocompromised patients can develop months-long infection with SARS-CoV-2, providing opportunities for within-host virus evolution and the emergence of immune escape mutations. Prior studies have identified immunologically important mutations in SARS-CoV-2 sequences from immunocompromised patients, particularly within the spike protein, which is required for entry and is the target of currently approved vaccines. For example, spike mutations in the angiotensin converting enzyme 2 (ACE2) receptor binding motif (RBM) such as E484K and Q493K^1,2^ and deletions in the spike N terminal domain (NTD)^3,4^ have been identified in immunocompromised patients with prolonged infection. These mutations have been found to confer partial resistance to neutralizing antibodies^5–9^ and are also are found in SARS-CoV-2 variants of concern. In part due to this, it has been hypothesized that prolonged SARS-CoV-2 infection in immunocompromised patients may contribute to the emergence of variants with global impact^10,11^.

However, immunologically important mutations do not arise in all immunocompromised patients with prolonged infection, and the lack of adaptive immune responses makes it possible that an additional selective pressure is required, such as exogenous antibody treatment. Understanding conditions that promote the emergence of immunologically important SARS-CoV-2 mutations in immunocompromised patients is critically important to both guiding treatment and potentially preventing the emergence of new SARS-CoV-2 variants. Here, we investigated the interplay between exogenous antibody treatment, endogenous humoral and cellular immune responses, and within-host virus evolution in five immunocompromised patients with prolonged SARS-CoV-2 infection.

## Results

### Overview of clinical courses

We identified five immunocompromised patients with persistent (> 30 days) SARS-CoV-2 infection (**Table 1, Figure 1**). All had a history of underlying malignancy and four were treated with immunosuppressive regimens including rituximab, while the other patient (P4) had Good Syndrome, resulting in hypogammaglobulinemia, B-cell deficiency, and CD4/CD8 T-cell imbalance. The duration of positive PCR tests for SARS-CoV-2 ranged from 42 to 302 days from the time of first positive test (d42-d302). Patients had persistently low SARS-CoV-2 PCR cycle threshold (C_T_) values throughout infection, as well as detectable subgenomic RNA (**Figure 1**). Two patients (P2 and P3) were treated with the single-agent monoclonal antibody (mAb) bamlanivimab soon after SARS-CoV-2 infection (d4 and d8, respectively). Two other patients were treated with high-titer convalescent plasma (CP): P4 at d0 and d104, and P5 at d196. Two patients (P3 and P4) received intravenous immunoglobulin (IVIG) as part of treatment for their underlying condition. All patients were treated with remdesivir and all but one (P1) were treated with steroids; there were no known changes to the patients’ baseline immunosuppressive regimens. All but one patient (P2) ultimately recovered.

**Table 1:**
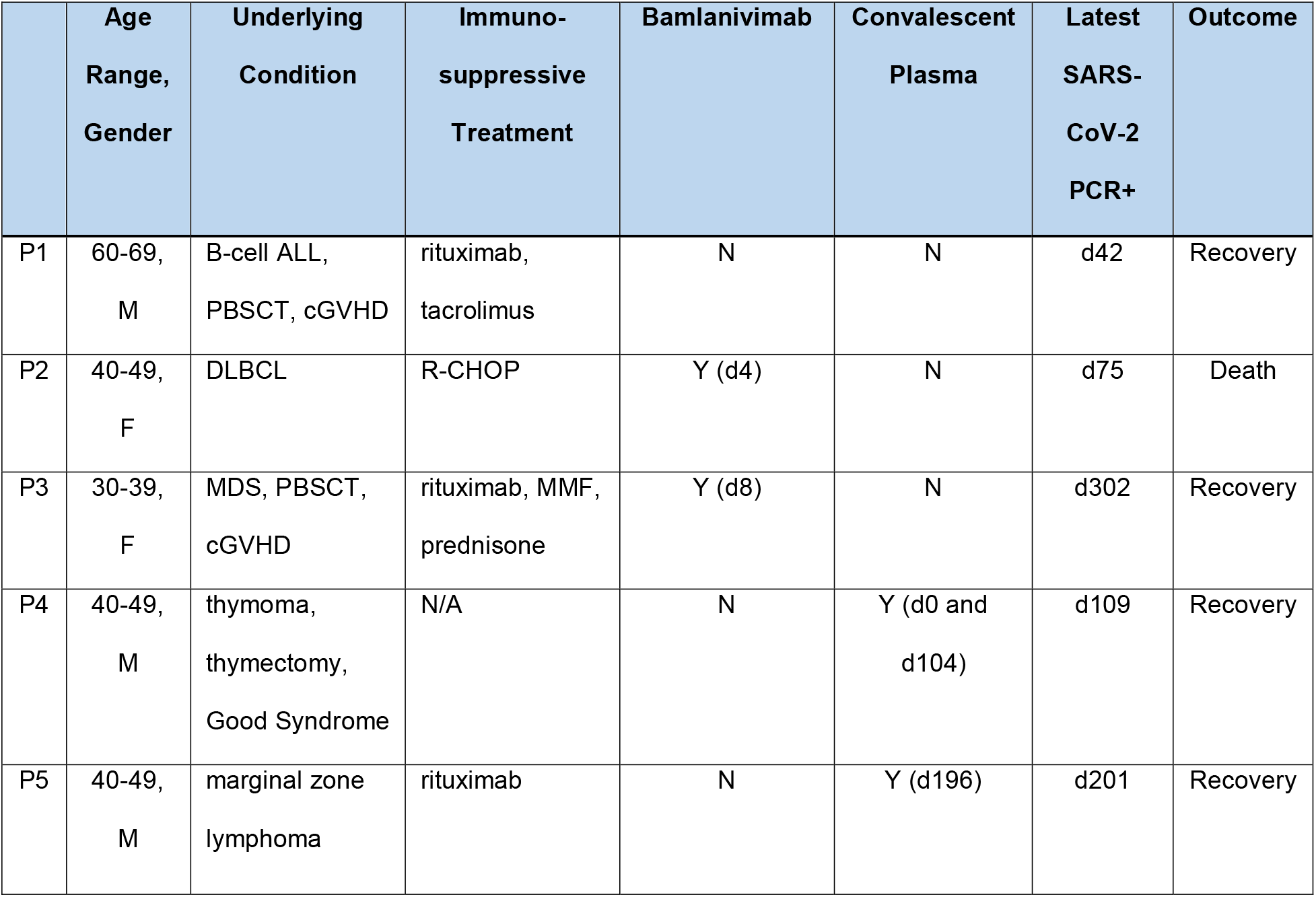
Clinical features of five immunocompromised patients with persistent SARS-CoV-2 infection. Abbreviations: ALL= acute lymphoblastic leukemia; cGVHD = chronic graft versus host disease; DLBCL = diffuse large B cell lymphoma; MDS = myelodysplastic syndrome; PBSCT = peripheral blood stem cell transplantation; R-CHOP = rituximab, cyclophosphamide, hydroxydanorubicin, vincristine sulfate, prednisone; MMF= mycophenolate mofetil; IVIG = intravenous immunoglobulin.

**Fig. 1:**
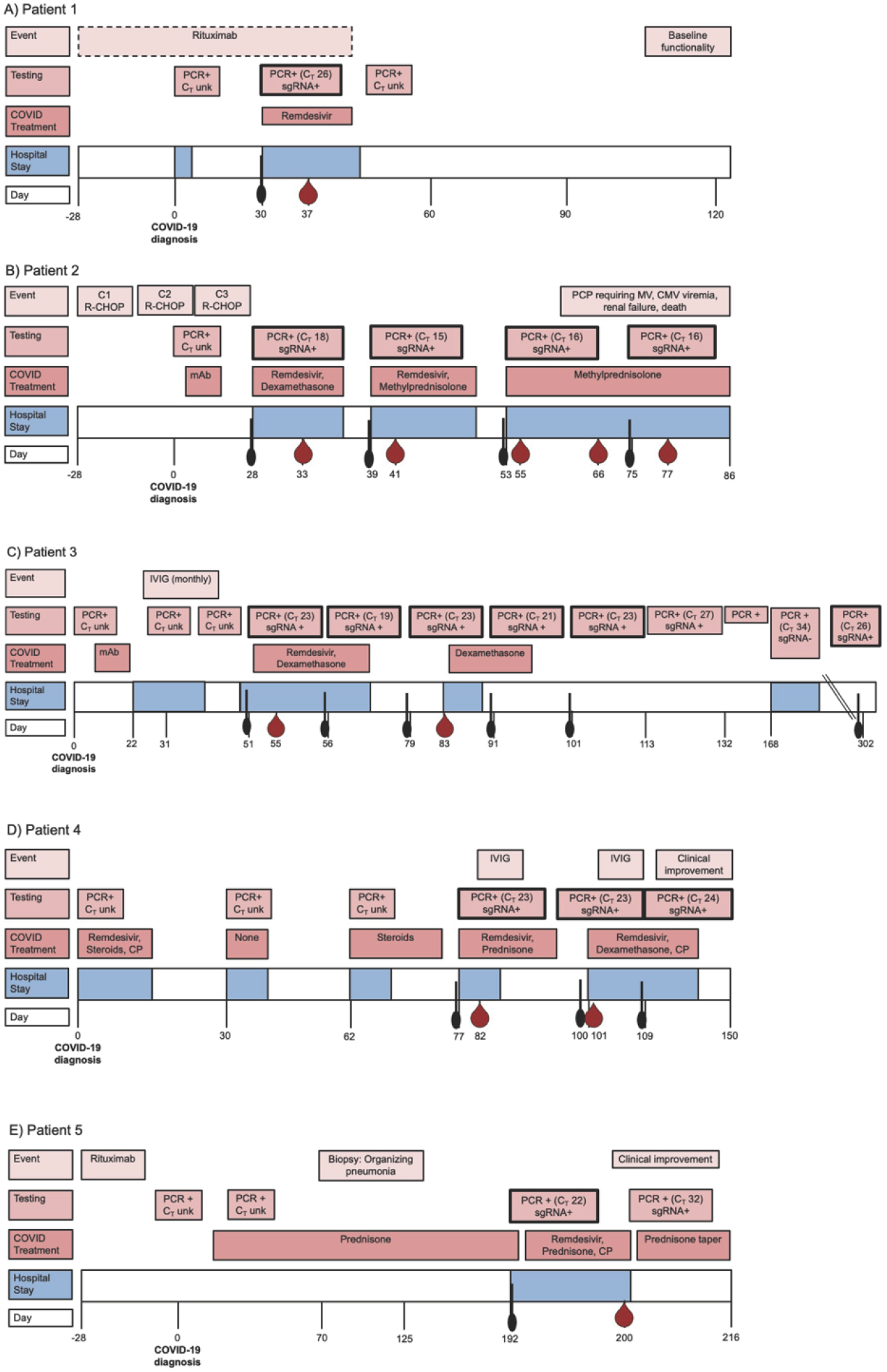
Five immunocompromised patients experienced prolonged SARS-CoV-2 infection despite multiple treatments. Panels indicate the timing of hospital admissions (blue), treatments (dark pink) and SARS-CoV-2 molecular testing (medium pink) for patients P1 (A), P2 (B), P3 (C), P4 (D), and P5 (E). Timeline is not to scale. C_T_ values shown are from confirmatory testing in the research laboratory to ensure consistency. Boxes with dark outlines and black swabs indicate nasopharyngeal samples used for SARS-CoV-2 sequencing, and blood drops indicate samples used for humoral or cellular analysis. Abbreviations: CMV = cytomegalovirus, CP = convalescent plasma, IVIG = intravenous immunoglobulin, R-CHOP = rituximab, cyclophosphamide, hydroxydanorubicin, vincristine sulfate, prednisone, mAb = monoclonal antibody (bamlanivimab), MV= mechanical ventilation, PCP = pneumocystis carinii pneumonia.

### Detailed clinical courses

#### Patient 1

This patient was a 60-69-year-old man with a history of relapsed B-cell acute lymphoblastic leukemia who underwent peripheral stem cell transplant (PBSCT) 18 months prior to his COVID-19 diagnosis. His post-PBSCT course was complicated by chronic graft-versus-host disease (GVHD) of the skin and gastrointestinal tract for which he was maintained on tacrolimus and rituximab infusions every 8 weeks. His last dose of rituximab was 9 days prior to his COVID-19 diagnosis, and he received rituximab again 47 days after his initial positive test. He initially presented with 3 days of nausea, vomiting, cough, and sore throat and was diagnosed with COVID-19 via nasopharyngeal (NP) PCR (cycle threshold [C_T_] unknown). He was not hypoxemic and chest X-ray showed no acute abnormalities. He was treated symptomatically and did not receive antivirals or undergo changes in immunosuppression, and he was discharged one day after admission. His symptoms initially resolved, but he developed recurrent cough and progressive shortness of breath and presented to the emergency department for further evaluation 29 days following his previous discharge. At that time, he was hypoxemic to 84% on room air, and chest CT showed peripheral areas of ground glass opacity and changes compatible with diffuse alveolar damage. His SARS-CoV-2 NP PCR was again positive, with C_T_ 26. This was felt to reflect ongoing COVID-19 infection, and he received a 5-day course of remdesivir. His symptoms improved over several days and he was discharged without supplemental oxygen 10 days after admission. He had no functional limitations at follow-up 3 months after the second hospitalization.

#### Patient 2

This patient was a 40-49-year-old woman diagnosed with stage IV diffuse large B-cell lymphoma 2 months prior to her COVID-19 infection, for which she had been treated with 2 cycles of R-CHOP; she last received rituximab 7 days prior to her COVID-19 diagnosis. Several days after the second cycle she developed cough and shortness of breath and was diagnosed with COVID-19 via NP PCR (C_T_ unknown). On day 4 of illness, she was treated with bamlanivimab and clinically improved, and she subsequently received cycle 3 of R-CHOP 17 days later. She then developed fever and progressive shortness of breath prompting readmission 7 days following cycle 3 of R-CHOP. SARS-CoV-2 NP PCR was again positive with C_T_ 18, and chest CT showed patchy groundglass opacities. IgG antibodies to the SARS-CoV-2 receptor binding domain were positive, and this was felt to reflect previous receipt of bamlanivimab. She was treated with 5 days of remdesivir and 10 days of dexamethasone. She was readmitted again one week later with worsening chills, shortness of breath, and hypoxemia. SARS-CoV-2 testing via NP PCR was again positive (C_T_ 15), and chest CT showed worsened opacities. She was treated with another 5-day course of remdesivir and steroid pulse then taper for possible organizing pneumonia. She was discharged after a 7 day hospitalization but was again admitted 7 days later for dyspnea and hypoxia to 86%. SARS-CoV-2 testing via NP PCR was positive (C_T_ 16), and chest CT showed progression of bilateral patchy opacities. She was continued on methylprednisolone, and her course was complicated by *Pneumocystis jirovecii* pneumonia despite prophylaxis, progressive respiratory failure requiring mechanical ventilation, CMV viremia, and renal failure. SARS-CoV-2 NP PCR obtained on day 22 of hospitalization was positive with a C_T_ 16, and she died on hospital day 33.

#### Patient 3

This patient was a 30-39-year-old woman with prior history of myelodysplastic syndrome who had undergone matched related PBSCT 3 years prior to her COVID-19 diagnosis. Her post-transplant course was complicated by chronic GVHD of the gastrointestinal tract, skin, and eyes, as well as CMV enteritis, and she was maintained on rituximab (last dose approximately 3 months prior to COVID-19 diagnosis), mycophenolate mofetil, prednisone, and monthly intravenous immunoglobulin infusions. She tested positive for COVID-19 via NP PCR (C_T_ unknown) after exposure to a known case; she had no symptoms at that time and received bamlanivimab 8 days later. Approximately 2 weeks after monoclonal antibody administration, she developed shortness of breath and hypoxia requiring hospital admission. A SARS-CoV-2 NP PCR was positive (C_T_ unknown) and chest CT showed multifocal groundglass opacities with areas of tree-in-bud nodularity. Given concern for pulmonary GVHD, she underwent bronchoscopy with transbronchial biopsy; BAL cultures were negative, and histology was unrevealing. She did not receive any treatment for COVID-19 during that admission, and no changes were made to her immunosuppression. One week following discharge, she was readmitted for worsening shortness of breath and fever. A repeat SARS-CoV-2 NP PCR was positive (C_T_ 23), and chest CT showed progression of patchy peripheral bilateral opacities. She was treated with 5 days of remdesivir and 10 days of dexamethasone. Repeat NP PCR after 5 days of remdesivir was positive with C_T_ 19. She was discharged, initially improved, but developed worsening shortness of breath after steroids were tapered, prompting readmission 3 weeks later. SARS-CoV-2 NP PCR was again positive (C_T_ 23), and chest CT showed new bilateral scattered groundglass opacities. Her clinical picture and radiographic findings were felt to reflect SARS-CoV-2 related inflammatory changes, and she was discharged home on a 4-week dexamethasone taper which was converted to a maintance dose of prednisone. She did well from a pulmonary standpoint, but was readmitted three months later and again seven months later, both times with CMV enteritis. During the first of those readmissions, at 260 days after her first PCR test, a clinical lymphocyte panel showed immune deficiencies in both T and B cell arms (534 CD3^+^ T cells/uL blood; 263 CD4^+^ T cells/uL blood; 294 CD8^+^ T cells/uL blood; 2 CD19^+^ B cells uL/blood). The patient’s SARS-CoV-2 NP PCR was positive on both admisssions (C_T_ 34 and 26), but she did not exhibit any respiratory symptoms. She continued to follow up with Infectious Diseases as an outpatient and eventually tested negative by home rapid antigen test 12 months after initial diagnosis.

#### Patient 4

This patient was a 40-49-year-old man with prior history of thymoma and subsequent thymectomy who developed cough, fever, and shortness of breath and was diagnosed with COVID-19 at another institution via NP PCR (C_T_ unknown). At that time, he was noted to be hypoxemic and was treated for COVID-19 with convalescent plasma, remdesivir, and steroids with clinical improvement. However, approximately 2 weeks later, his symptoms worsened and he was readmitted to the same facility. At that time, he was was presumptively treated for bacterial pneumonia with antibiotics but did not receive any dedicated treatment for COVID-19, and he was discharged on supplemental oxygen. Approximately 2 weeks later, he experienced recrudescence of fever, chills, cough, and shortness of breath and was hospitalized at the same facility; he was treated with antibiotics for possible bacterial pneumonia and was discharged on a steroid taper. He was subsequently admitted to our facility given lack of improvement. At that time, SARS-CoV-2 NP PCR was positive (C_T_ 23), and extensive testing for secondary bacterial and fungal infections was negative. Given concern for persistent SARS-CoV-2 infection, he received 10 days of remdesivir and dexamethasone. His laboratory evaluation was otherwise significant for hypogammaglobulinemia, B-cell dysregulation, an abnormal CD4/CD8 ratio, and lack of lymphocyte response to tetanus and *Candida* antigens. In the context of these findings and prior thymoma, he was diagnosed with Good Syndrome and was treated with IVIG. Approximately 3 weeks later, he was again readmitted to our center with acute hypoxic respiratory failure due to COVID-19. SARS-CoV-2 NP PCR was positive (C_T_ 23). At that time, he was treated with 10 days of remdesivir and dexamethasone and received two doses of convalescent plasma for the management of SARS-CoV-2 infection; he also received a dose of IVIG for Good Syndrome. Convalescent plasma was found to be high titer either based on signal-to-cutoff (S/C) ratio ≥ 9.5 on the VITROS Anti-SARSCoV-2 IgG assay (Ortho Clinical Diagnostics, Raritan, NJ) or a cut off index ≥ 109 or a titer of ≥ 132 U/mL on the Elecsys Anti-SARSCoV-2 assay (Roche Diagnostics International Ltd, Rotkreuz, Switzerland). He was seen in follow up approximately 3.5 months following discharge; at that time, he was doing well and continued on monthly IVIG for his underlying immunodeficiency state.

#### Patient 5

This patient was a 40-49-year-old man with marginal zone lymphoma diagnosed approximately 3 years before developing COVID-19. At the time of his lymphoma diagnosis, he was treated with bendamustine and rituximab and achieved remission; thereafter, he was continued on monthly maintenance rituximab for approximately 2 years. One month after stopping rituximab, he developed cough and shortness of breath and was diagnosed with COVID-19; the details surrounding his initial diagnosis and treatment are unknown. He continued to have dyspnea on exertion and exertional hypoxemia three months following his COVID-19 diagnosis. He underwent video-assisted thoracoscopic biopsy, and pathology showed changes compatible with organizing pneumonia secondary to COVID-19; there was no evidence of superimposed infectious or malignant process. He was treated with prednisone (40mg daily), but did not have any improvement in symptoms over the next 4 months. In this context, he was admitted to our facility for ongoing management. At that time, SARS-CoV-2 NP PCR was positive (C_T_ 22), and chest CT showed patchy multifocal and confluent groundglass opacities. Given concern for persistent COVID-19 infection in the setting of B-cell depleting therapy, he received a 5-day course of remdesivir and a dose of convalescent plasma. Convalescent plasma was found to be high titer either based on signal-to-cutoff (S/C) ratio ≥ 9.5 on the VITROS Anti-SARSCoV-2 IgG assay (Ortho Clinical Diagnostics, Raritan, NJ) or a cut off index ≥ 109 or a titer of ≥ 132 U/mL on the Elecsys Anti-SARSCoV-2 assay (Roche Diagnostics International Ltd, Rotkreuz, Switzerland). A subsequent SARS-CoV-2 NP PCR obtained 8 days following admission and completion of SARS-CoV-2-directed therapies was again positive (C_T_ 32). He improved symptomatically and was discharged home. He was seen in outpatient follow up approximately 2.5 months later and had remained off steroids and supplemental oxygen.

### Immune responses

Multiple immune measurements underscored the impact of exogenous antibody treatment and the lack of an endogenous antibody response in these patients. Both patients who received bamlanivimab within 8 days (P2 and P3) had high serum IgG titers to a pre-fusion stabilized spike trimer of reference virus Wuhan-Hu-1 and potent pseudovirus neutralizing serum titers to the reference virus at the earliest time points tested (d33 and d55, respectively) (**Figure 2**). They retained elevated, though decreasing, anti-spike IgG and neutralizing antibody titers through the last time points tested (d77 and d83, respectively). Of the two patients who received CP, P5 had low but detectable anti-spike IgG and neutralizing antibody titers at d200, four days after receiving CP. By contrast, P4, who received CP at the onset of infection, did not have detectable anti-spike IgG or detectable neutralizing antibody titers to the reference virus at d82 and d101. P1 did not receive exogenous antibody treatment, and anti-spike IgG and neutralizing antibody titers were negative at d37, indicating a lack of endogenous immune response.

**Fig. 2:**
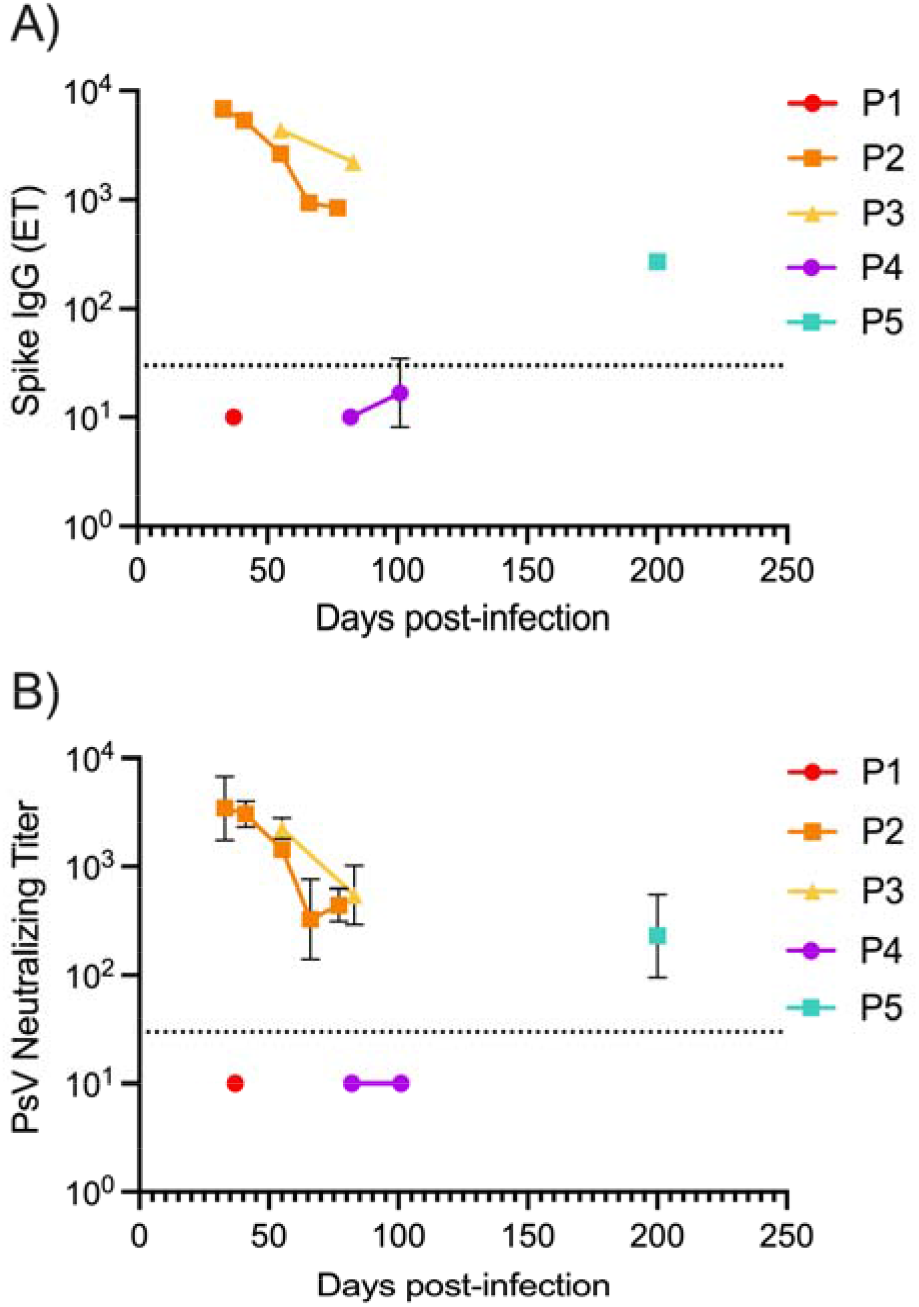
Antibody responses to SARS-CoV-2 reference isolate in immunocompromised patients reflect exogenous antibody treatments. **A)** Endpoint IgG titers to SARS-CoV-2 Wuhan-Hu-1 spike trimer in serum samples collected from immunocompromised patients at various time points post-infection. **B)** Neutralizing titers of patient sera against a SARS-CoV-2 Wuhan-Hu-1 pseudovirus (psV) at various time points post-infection. PsV neutralizing titers represent the reciprocal serum dilution at which half-maximal psV neutralization was observed, or IC50. Data show geometric means and geometric SD from 2-5 independent experiments. The dotted line represents the limit of detection (LOD).

We examined the peripheral cellular immune compartment for the three immunocompromised patients for whom whole blood samples were available. Immunocompromised patients P2, P4, and P5 had lower frequencies of lymphocytes within peripheral blood mononuclear cells (PBMC; 2.4% for P2; 62.3% ad 38.7% for P4 at d82 and d101; 24.0% for P5) compared to healthy controls (59.9% and 68.4%) (**Figure 3**). Age-matched patients hospitalized with COVID-19 also had low frequencies of lymphocytes within PBMC (48.3% and 31.1%) (**Figure 3**), consistent with clinical lymphopenia described with COVID-19^12,13^.

**Fig.3:**
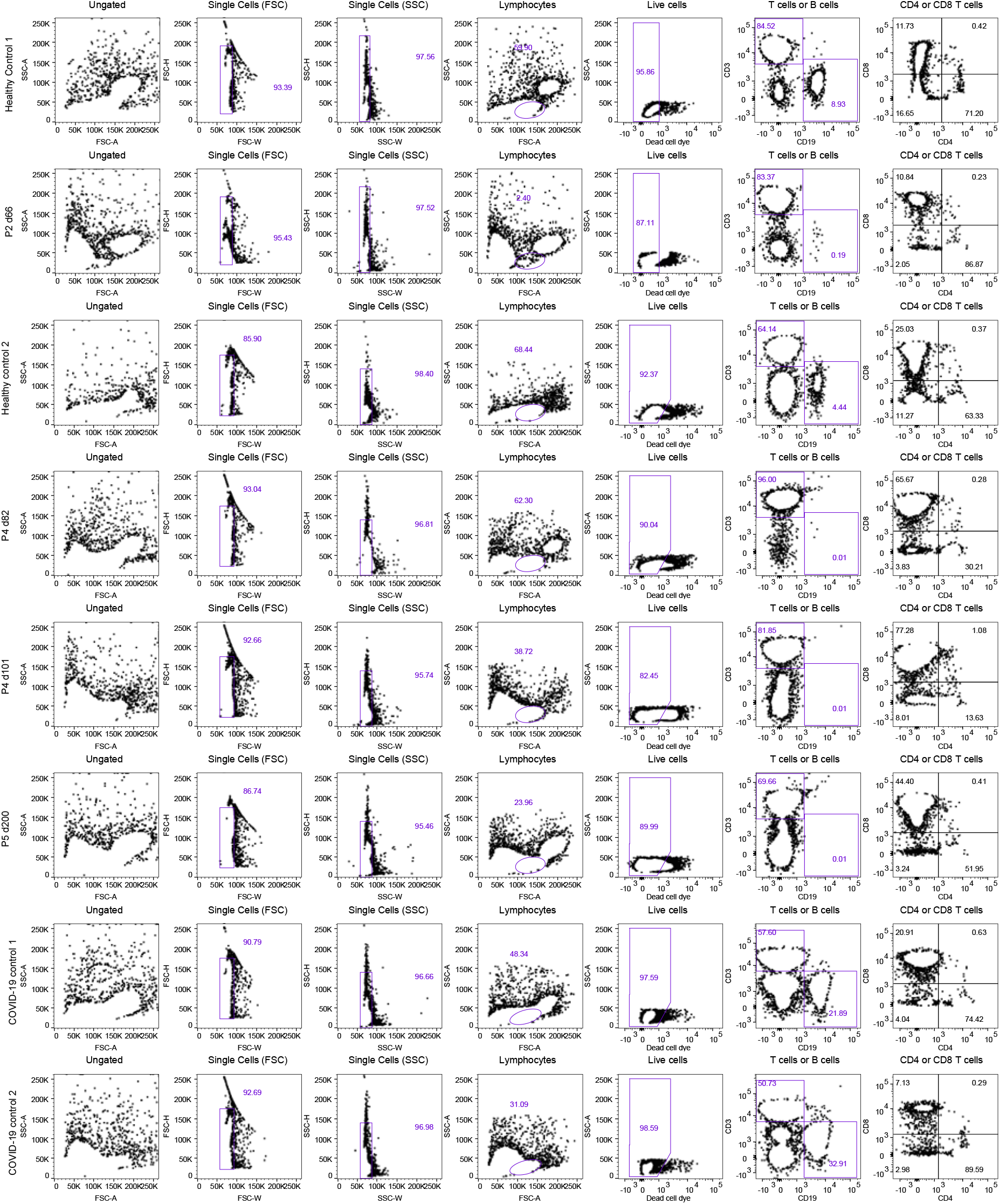
Gating strategy used to identify patient frequencies of lymphocytes, B cells, and T cells. PBMC from two healthy control subjects, immunocompromised patients P4 (d82 and d101) and P5 (d200), and two age-matched patients hospitalized with COVID-19 (COVID-19 controls 1 and 2) were stained with a dye that penetrates dead cells and fluorescent antibodies against surface markers and analyzed by flow cytometry. B cells and T cells are identified by the lineage specific markers CD19 or CD3, respectively.

The three immunocompromised patients had low to undetectable frequencies of CD19^+^ B cells within the lymphocyte population (0.19% for P2, 0.01% for P4 at d82 and d101, and 0.01% for P5) compared to healthy controls (8.93% and 4.44%) or COVID-19 controls (21.89% and 32.91%) (**Figure 3**). We were not able to measure cellular responses for P3 due to severe anemia, however the patient received rituximab 3 months prior to infection, and at d260 still exhibited clinically low T and B cell counts. Thus, basic immune phenotyping data suggest that the antibody responses against reference SARS-CoV-2 virus observed in P2, P3, and P5 were due to exogenous treatment rather than an endogenous immune response.

Although patients P2, P4, and P5 all had CD3^+^, CD4^+^, and CD8^+^ T cells, only P4 and P5 had robust SARS-CoV-2 specific T cell responses to SARS-CoV-2 peptide pools (**Figure 4**). This response was predominantly effector CD8^+^ T cells secreting antiviral IFNg or both IFNg and TNF following *ex vivo* PBMC stimulation with pools containing spike peptides, but also included multi-functional CD4^+^ T cell responses against pools containing spike peptides for P5. Both P4 at d82 and d101 and P5 at d200 exhibited higher magnitude SARS-CoV-2 CD8^+^ T cells responses than either age-matched COVID-19 control at d13 or d18. In contrast, P2 at d66 and the healthy controls had only baseline levels of SARS-CoV-2 specific T cell responses (**Figure 5, Figure 6**). All subjects elicited T cell responses to a positive control antigen.

**Fig. 5:**
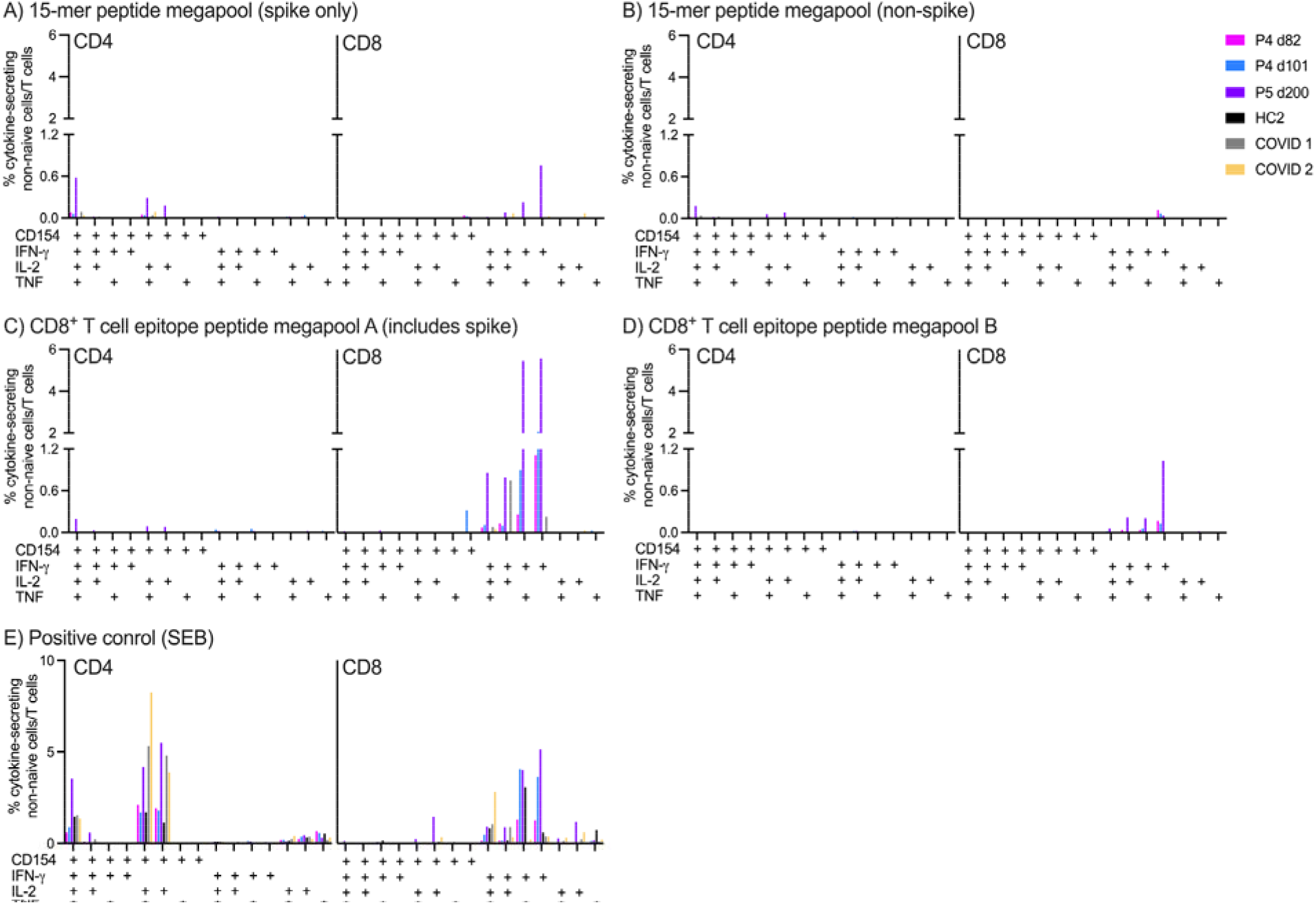
Patients P4 and P5 elicit functional CD8 and CD4 T cell responses against SARS-CoV-2. PBMC isolated from patient P4 at d82 and d101 and from patient P5 at d200, as well as from a healthy control donor (HC2) and two age-matched patients hospitalized with COVID-19 (COVID 1 and 2) were stimulated with a peptide megapool containing A) 15-mers from the spike ORF, B) 15-mers from all other non-spike SARS-CoV-2 ORFs, C) predicted CD8^+^ T cell epitopes from ORFs, including spike, or D) predicted CD8^+^ T cell epitopes from non-spike ORFs; or E) a positive control antigen (*Staphylococcus enterotoxin* B, SEB); or a negative control (media with equivalent peptide vehicle). Frequencies of non-naive T cells out of total CD154, IFN-*γ*, TNF, or IL-2 expressing CD4^+^ or CD8^+^ T cells responding to each stimulation were measured by flow cytometry, and background responses subtracted.

**Fig. 6:**
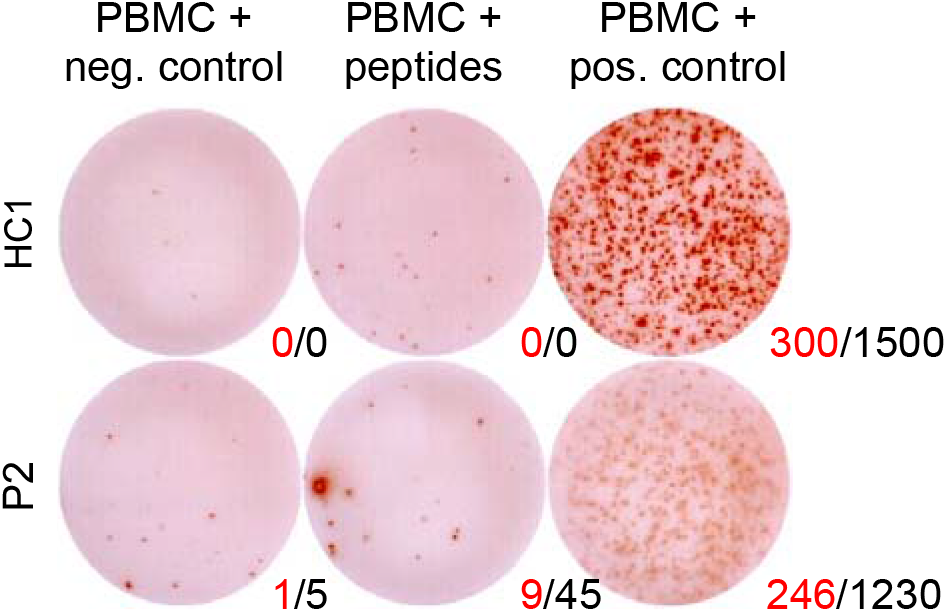
Patient P2 elicits background levels of SARS-CoV-2-specific T cell responses. PBMC from patient P2 at d66 and from a healthy control (HC1) were separately stimulated with a peptide megapool containing 15-mers against all SARS-CoV-2 ORFs, a negative control (media with equivalent peptide vehicle), or a positive control (staphylococcal enterotoxin B, a superantigen). IFN-*γ*-secreting cells were detected by ELISPOT and quantified as spot-forming units (SFU) per well (numbers in red) and SFU per million PBMC (number in black). The assay limit of detection is <10 SFU/well.

### Intra-patient SARS-CoV-2 evolution

To assess within-patient SARS-CoV-2 diversity and evolution, we performed high-depth viral sequencing from residual nasopharyngeal samples collected across 1-6 time points per patient (**Figure 1**). Phylogenetic analysis of consensus SARS-CoV-2 sequences indicated that patients were infected with viral lineages circulating in the community (**Figure 7**). Sequences belonged to SARS-CoV-2 lineages B.1.2 (P1, P2, and P3), B.1.568 (P4), and B.1.493 (P5). Longitudinal samples were available from patients P2, P3, and P4, and consensus sequences from each were monophyletic, reflecting within-host evolution. Compared to the first available time point, between 4 and 26 consensus single nucleotide polymorphisms (SNPs) arose within each patient, most of which were nonsynonymous and many of which occurred in the spike protein **(Figure 8)**.

**Fig. 7:**
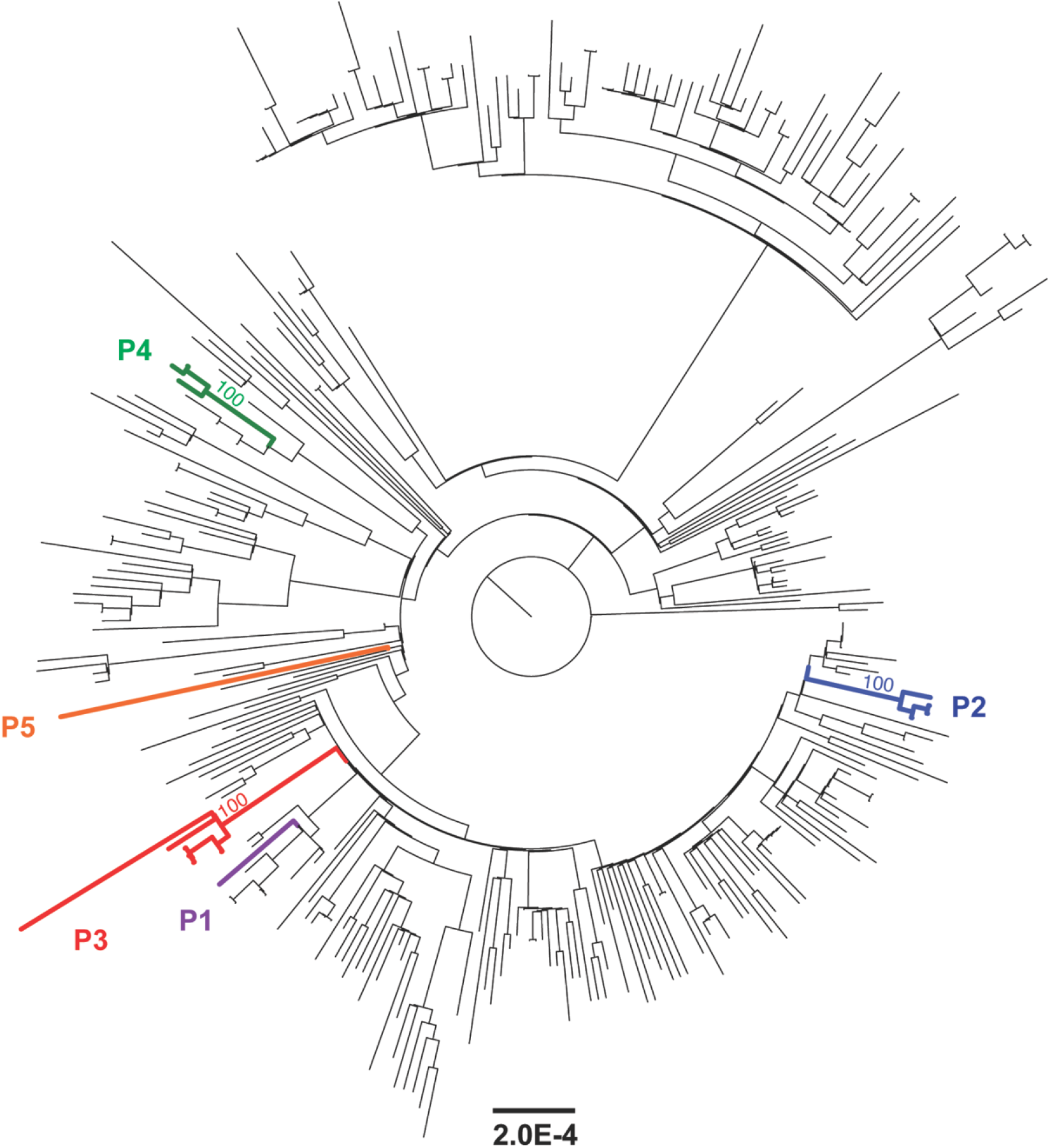
SARS-CoV-2 consensus sequences from each immunocompromised patient indicate infection from the community and no evidence for reinfection in patients with longitudinal samples. Maximum-likelihood tree includes consensus SARS-CoV-2 sequences from P1 (n = 1), P2 (n = 4), P3 (n = 6), P4 (n = 3), and P5 (n = 1), as well as 301 reference sequences from patients within the Emory Healthcare system between 1/1/2021 and 4/30/2021. Sequences from each of the three patients with longitudinal samples form monophyletic clades, indicating no evidence for reinfection. 1,000 bootstrap replicates were performed, and percent bootstrap support is shown for the most recent common ancestor of each immunocompromised patient with longitudinal sampling.

**Fig. 8:**
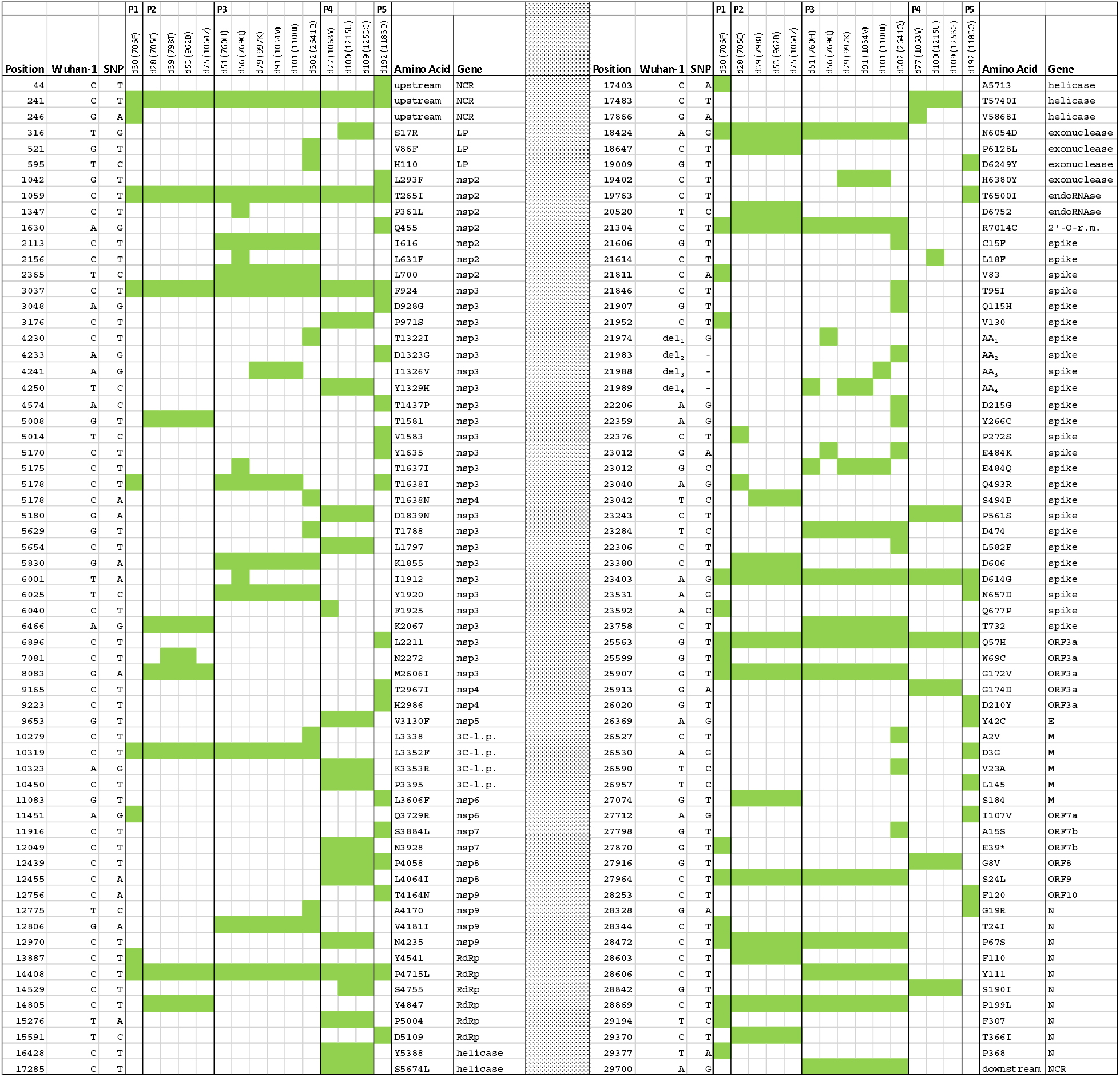
Consensus-level SNPs across patients. Filled green squares show SNPs that reach the consensus level relative to the Wuhan-Hu-1 reference, annotated by gene and corresponding amino acid change (if applicable). Sites where within-patient consensus-level changes occur over the course of infection are ones that transition from green to unfilled squares or from unfilled squares to green squares. Abbreviations: non-coding region (NCR); leader protein (LP); nonstructural protein (nsp); RNA-dependent RNA polymerase (RdRp); 3C-like proteinase (3C-l.p.); 3’-to-5’ exonuclease (exonuclease); 2’-O-ribose methyltransferase (2’-O-r.m.); ORF3a protein (ORF3a); envelope (E); membrane glycoprotein (M); ORF7a protein (ORF7a); ORF8 protein (ORF8); nucleocapsid phosphoprotein (NcPh); deletion from A21974 to 21994 (del_1_); deletion from D138-Y144 and Y145D (AA_1_); deletion from T21983 to 21994 (del_2_); deletion from L141 to Y144 (AA_2_); deletion from T21986 to 21994 (del_3_); deletion from G142-Y144 and Y145D (AA_3_); deletion from T2189 to 21994 (del_4_); deletion from V143-Y144 and Y145D (AA_4_).

Through analysis of deep sequencing data, we identified intra-sample single nucleotide variants (iSNVs) present at >2% frequency in the patients with longitudinal samples available (P2-P4). We identified both iSNVs and fixed mutations in spike regions known to be associated with immune escape (**Figure 9**). P2 and P3 (treated with bamlanivimab) experienced rapid evolution in the spike RBM, the target of bamlanivimab. Specifically, in P2, RBM substitution Q493R (compared to Wuhan-Hu-1) was present at d28 but reverted to the ancestral Q493 just 11 days later, at d39. At the same time, substitution S494P arose at the adjacent amino acid position. Both sites remained monomorphic until d75, when Q493R and S494 were again observed at frequencies of 20-25%. RBM substitution E484K also arose in P2 between d28 and d39 and remained detectable thereafter, but only at intermediate frequencies (29-45%), with the ancestral E484 remaining dominant. Evolution at site 484 also occurred in P3, in whom E484Q was fixed at d51, transiently became polymorphic with E484K at d56 (66% frequency E484K, 34% frequency E484Q), reverted to fixed with E484Q at d79, d91, and d101, and was fixed with E484K at d302. Thus, in both P2 and P3, substitution E484K was present either intermittently or at intermediate frequency.

**Fig. 9:**
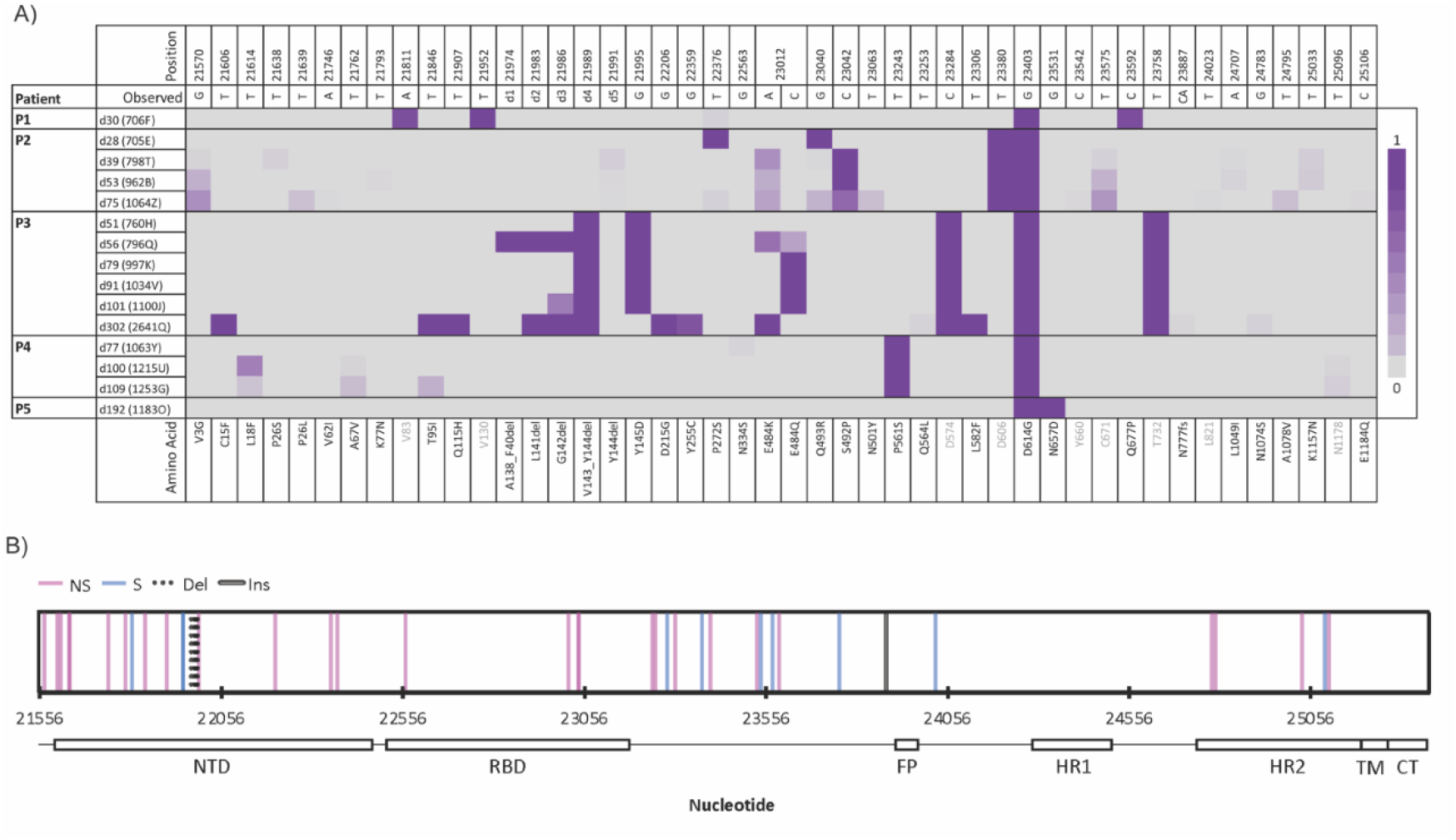
SARS-CoV-2 spike mutations from five immunocompromised patients. Mutations in the SARS-CoV-2 spike gene for each patient and time point, compared to Wuhan-Hu-1. Shading denotes mutation frequency. For each mutation, the variant nucleotide listed in the ‘Observed’ row, and the amino acid mutation is listed below the plot. Gray text indicates synonymous mutations. Abbreviations: fs = frameshift, del = amino acid deletion, d1 = deletion from 21974-82; d2 = deletion from 21983-5; d3 = deletion from 21986-8; d4 = deletion from 21989-94; d5 = deletion from 21991-3. B) Mutations in the SARS-CoV-2 spike gene for all patients and time points, mapped to their locations on the genome. Abbreviations: NS = nonsynonymous, S = synonymous, Del = deletion, Ins = insertion, NTD = N terminal domain, RBD = receptor binding domain, FP = fusion peptide, HR1 = heptad repeat 1, HR2 = heptad repeat 2, TM = transmembrane, CT = C-terminal.

We also observed changes in the spike NTD, a target of some potently neutralizing antibodies^4^ but not bamlanivimab (**Figure 9**). In P2, a proline residue rapidly replaced the serine residue at site 272 between d28 and d39, with a low level of serine circulation apparent again at d75. In P3, NTD deletion V143-Y144 was present throughout infection, accompanied by adjacent substitution Y145D at all time points but the last. In addition, there were transient deletions of the upstream 5 amino acids (138-142) at d56, the upstream 1 amino acid (142) at d101, and the upstream 2 amino acids (141-142) at d302. These deletions occurred in the NTD recurrent deletion region 2 (RDR2)^4^, where similar deletions have been observed in other immunocompromised patients^1,2,14–16^.

In contrast to the virus evolution observed in two patients treated with bamlanivimab, we did not detect consensus-level mutations or iSNVs in the RBM or NTD for P4 at either d77, d100, or d109 (**Figure 8, Figure 9**), after CP treatments at d0 and d104. For each of the other two patients, only one sample was available. P1, who was not treated with exogenous antibodies, did not have consensus-level spike RMB or NTD mutations at d30, but did have substitution Q677P in the spike S1 subunit. P5 was infected for substantially longer and did not have consensus-level spike RBM or NTD mutations prior to treatment with CP at d192. Only one iSNV was observed in P1, and none in P5, which may have been due to low sequencing depth (**Table 2**). Overall, we thus observed the emergence of spike mutations associated with immune evasion in only the patients undergoing bamlanivimab treatment.

**Table 2:** Sequencing metrics. Total reads indicates total number of metagenomic sequencing reads obtained per sample, and is the sum of at least two independent sequencing libraries. Coverage indicates percent SARS-CoV-2 genome coverage based on reference-based assembly to NC_045512.1, and mean depth indicates mean depth of coverage at each SARS-CoV-2 nucleotide position.

| **Sample** | **Total Reads** | **Coverage** | **Mean depth** |
| --- | --- | --- | --- |
| **Patient 1** |  |  |  |
| d30: 706F | 170,162,696 | 100% | 141 |
| **Patient 2** |  |  |  |
| d28: 705E | 17,361,712 | 100% | 4,656 |
| d39: 798T | 20,598,872 | 100% | 12,303 |
| d53: 962B | 17,485,750 | 100% | 7,907 |
| d75: 1064Z | 15,756,412 | 100% | 12,555 |
| **Patient 3** |  |  |  |
| d51: 760H | 326,861,238 | 100% | 427 |
| d56: 769Q | 19,928,108 | 100% | 1,589 |
| d79: 997K | 373,163,364 | 100% | 641 |
| d91: 1034V | 519,308,996 | 100% | 3,031 |
| d101: 1100J | 499,743,018 | 100% | 432 |
| d302: 2641Q | 239,745,572 | 100% | 210 |
| **Patient 4** |  |  |  |
| d77: 1063Y | 376,481,746 | 100% | 352 |
| d100: 1215U | 353,338,568 | 100% | 386 |
| d109: 1253G | 188,162,712 | 100% | 358 |
| **Patient 5** |  |  |  |
| d192: 1183O | 105,667,508 | 100% | 185 |

### Neutralization of autologous spike variants

To assess whether the variant spike proteins from P1, P2, and P3 were recognized by antibodies in autologous serum, we constructed SARS-CoV-2 pseudoviruses using replication incompetent lentiviruses expressing spike from the reference virus Wuhan-Hu-1, as well as the spike variants. All pseudoviruses infected a human cell line expressing ACE2 and could be neutralized by the human-derived anti-SARS-CoV-2 mAb CC12.1^17^ (**Figure 10**).

**Fig. 10:**
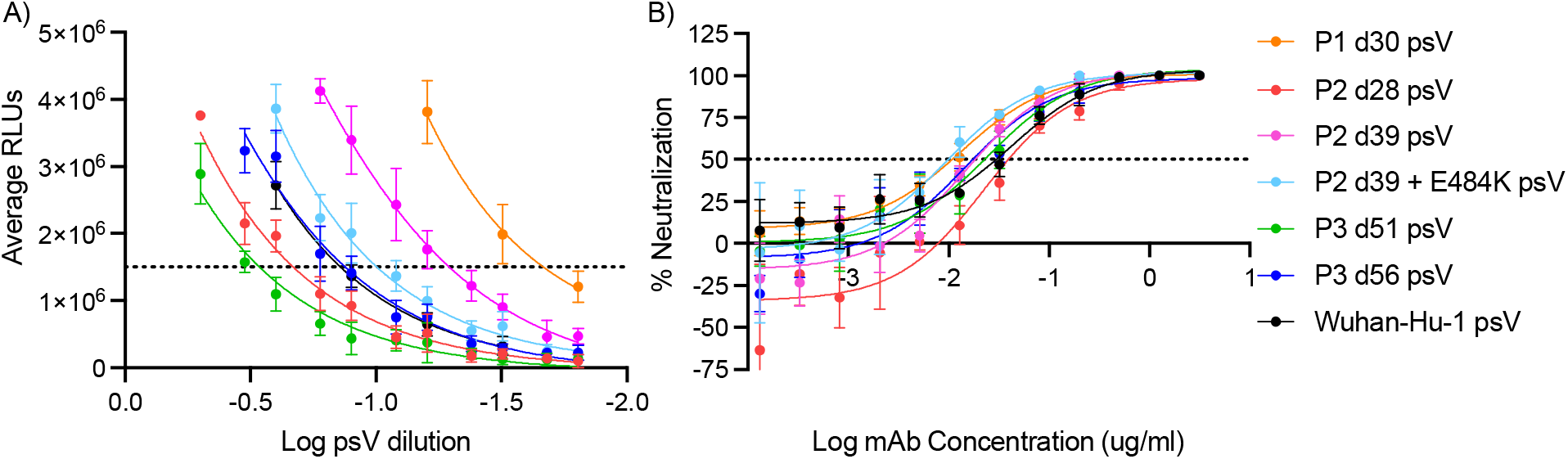
Autologous variant pseudoviruses (psV) are infectious and neutralized by mAb CC12.1. A) Serial dilutions of transfection supernatants containing indicated psV were incubated with ACE2-expressing cells and assessed for expression of a luciferase reporter plasmid as a measure of productive infection. All psV were infectious, as demonstrated by high levels of background-subtracted luminescence, quantified in relative light units (RLUs), on the y-axis. Variation in RLUs obtained for different psV is not necessarily indicative of differences in psV infectivity, as it could also reflect variation in transfection efficiencies. B) Graphs show the ability of mAb CC12.1 to neutralize SARS-CoV-2 psV with Wuhan-Hu-1 spike or spikes expressing mutations corresponding to autologous viral variants P1 d30, P2 d28, P2 d39, P2 d39 + E484K (P2 minor variant), P3 d51, or P3 d56.

Sera from patient P1, who did not receive exogenous antibody treatment, could not neutralize either pseudovirus containing autologous spike protein from d30, or reference virus, suggesting this patient did not elicit SARS-CoV-2 neutralizing antibodies. In contrast, sera or plasma from P2 and P3 neutralized the reference virus, but not pseudovirus containing autologous spikes (**Figure 11**). Specifically, five P2 samples from d33-d77 were unable to neutralize pseudovirus with spike from either d28 (containing P272S and Q493R) – or d39 (containing S494P, with or without E484K). Two P3 samples from d55 and d83 were unable to neutralize pseudovirus with the spike from either d51 (containing VY143del, Y145D, and E484Q) or d56 (containing DPFLGVY138del, Y145D, and E484K). These results suggest that, in P2 and P3 (treated with bamlanivimab), spike mutations conferring resistance to neutralization by serum containing active mAb had already emerged by the first sampled time point (d28 and d51, respectively). Moreover, the virus remained resistant to serum neutralization after continued spike evolution.

**Fig. 11:**
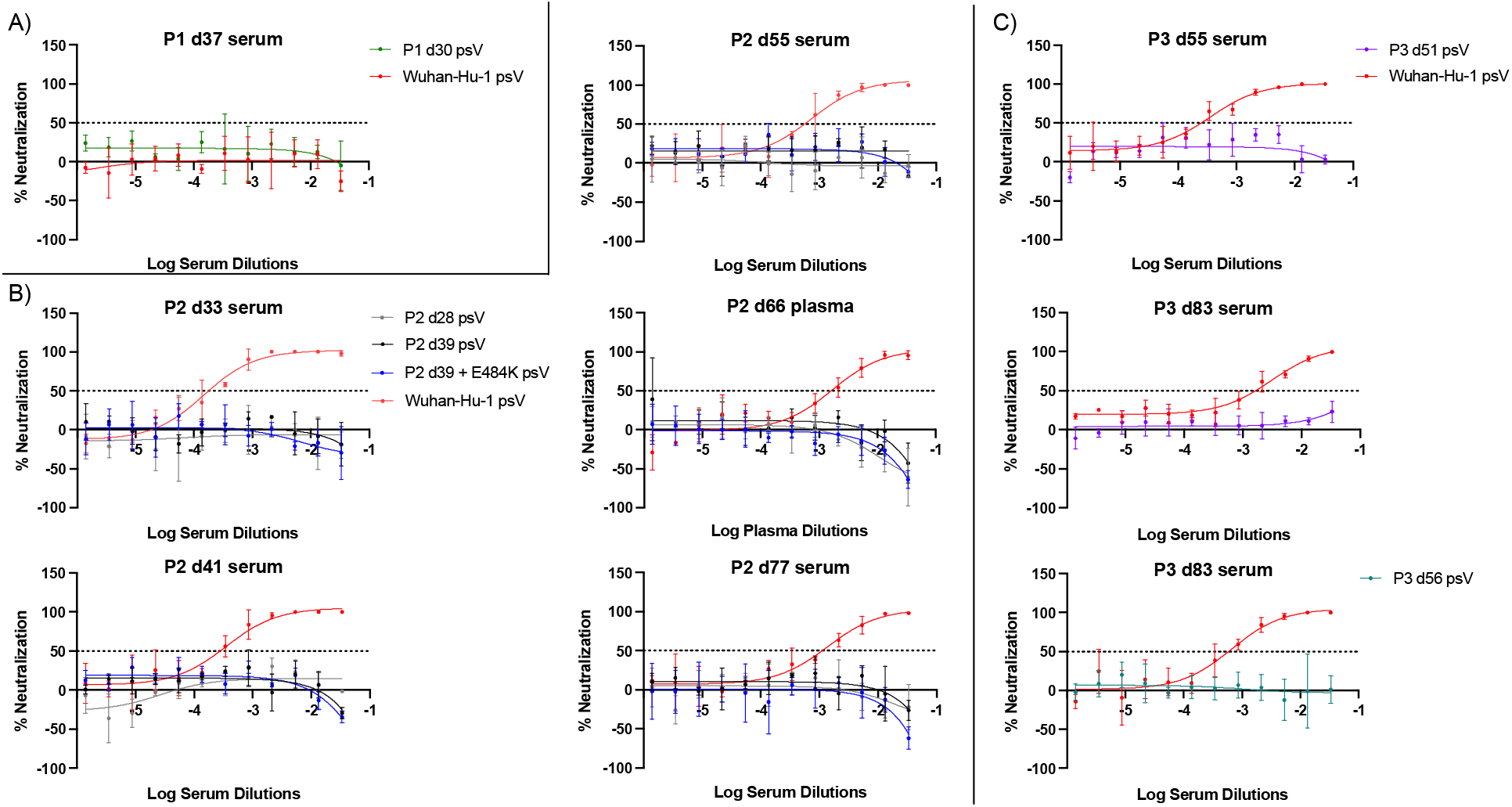
Sera/plasma from patients in immunocompromised cohort cannot neutralize autologous viral variants. Graphs show the ability of sera/plasma from patients P1 (A), P2 (B), or P3 (C) at indicated time points post infection to neutralize SARS-CoV-2 pseudoviruses with Wuhan-Hu-1 spike or spikes expressing mutations corresponding to autologous viral variants P1 d30, P2 d28, P2 d39, P2 d39 + E484K (P2 minor variant), P3 d51, or P3 d56. A serum, plasma, or monoclonal antibody is designated non-neutralizing if it does not achieve 50% neutralization at the highest concentrations tested. Graphs represent the mean and SD of three replicates and are representative of at least two independent experiments.

## Discussion

Our results underscore the complex interplay between antibody-based treatment, endogenous immune responses, and within-host SARS-CoV-2 evolution in immunocompromised patients with prolonged SARS-CoV-2 infection. By examining viral evolution and both humoral and cellular immune responses from multiple patients, our study represents one of the most comprehensive investigations to date.

Overall, our results suggest that persistent infection itself may not be sufficient to promote the emergence of immunologically relevant spike mutations. Instead, opportunities for immune escape arise when persistent virus replication is combined with selective pressure such as single-agent mAb treatment and the absence of effective endogenous immune responses. We detected viruses with neutralization resistant spike mutations 1-2 months after treatment with bamlanivimab, suggesting that escape from neutralization by exogenously supplied antibody may be a key factor in allowing persistent infection, and supporting the possibility that mAbs may contribute to the emergence of resistance mutations at the population level. Due to concerns about the emergence of resistance mutations, the FDA emergency use authorization for single-agent bamlanivimab therapy was revoked, and patients have subsequently been treated with alternative single or multi-agent mAb therapies. Multi-agent mAb therapy is theoretically less likely to select for neutralization resistance^18^, though it has been reported^19,20^. Although some previous studies have indicated that convalescent plasma may also exert selective pressure during longitudinal SARS-CoV-2 infection^3,15,16^, others have not^21^, and we did not observe escape mutations in the two patients who were treated with CP. This is potentially due to the timing of treatment compared to sample collection, or a lower effective antibody titer of CP compared to mAb therapy.

Based on the rapid emergence of spike mutations that did not confer changes in neutralization susceptibility, we propose that selective pressures other than single-agent bamlanivimab therapy were also at play in our patients with longitudinal infection. For example, in P2, the original spike variant with RBM substitution Q493R rapidly reverted to Q493, while over the same time, replacement with S494P occurred at the adjacent site. Both variants were neutralization resistant, consistent with prior reports^22,23^. Because the Q493/S494P variant rapidly rose in frequency and persisted throughout infection, it likely had a fitness advantage compared to Q493R/S494. One plausible explanation is ACE2 affinity, since S494P has similar binding compared to wild-type virus^23,24^, whereas Q493R is predicted to have weaker binding^25^. Supporting the higher fitness of S494P, it is much more common at a population level than Q493R, with 12,664 sequences^26^ compared to 252 sequences^27^ worldwide as of 2021-11-19. Underscoring the importance of complex fitness tradeoffs, spike RBM mutation E484K was detected at sub-consensus levels in P2 and transiently in P3. In both patients, E484K arose in the setting of existing neutralization resistance and did not alter the virus’ neutralization profile. It may not have achieved fixation due to potentially weaker binding to ACE2^24^, but its persistence at intermediate frequency could suggest cooperative interactions between variants^28^.

Finally, we observed deletions of fluctuating length in the spike NTD RDR2^4^ in one patient (P3). NTD deletions are most often reported in patients treated with CP^2,14–16^, unlike our patient, but they also have been reported in immunocompromised patients without exogenous antibody treatment^29^, as well as in publicly available sequences, not all of which are likely to be from immunocompromised patients^4^, and in one immunocompetent patient^30^. Similar to our results, prior studies have observed NTD and RBM mutations arising as minor variants^2^ and/or transiently^1^. Overall therefore, it is apparent that SARS-CoV-2 has multiple pathways available to achieve neutralization resistance, and additional selective pressures are important in determining within-host evolution in patients with longitudinal infection.

Interestingly, both patients in whom we did not observe spike RBM or NTD mutations (P4 and P5) were treated with CP and had SARS-CoV-2-specific T cell responses. P4 and P5 clinically improved soon after CP treatment, however larger studies have not demonstrated a benefit to CP, and it is no longer recommended for treatment^31^. Thus, our results raise interesting questions about the role of CD8^+^ T cells in immunocompromised patients. First, our results confirm that patients with B cell deficiencies can elicit effector T cells^32^. Other studies have observed functional SARS-CoV-2-specific CD4^+^ and CD8^+^ T cell responses in patients with B cell deficiencies, including higher magnitude CD8^+^ than CD4^+^ T cell responses^1,32,33^. However, those studies were smaller or did not also examine humoral responses. Second, our results suggest that CD8^+^ T cells may be critical to the resolution of SARS-CoV-2 infection, as suggested by other studies of immunocompromised patients^34^. Specifically, we observed clinical recovery in P4, who had no detectable neutralizing antibody response but did have a functional SARS-CoV-2-specific CD8^+^ T cell response, and in P5, who had functional SARS-CoV-2 specific CD4^+^ and CD8^+^T cell responses. By contrast, P2 succumbed to disease, after exhibiting no detectable neutralizing antibody response to autologous virus and only baseline SARS-CoV-2-specific T cell responses.

Notably, both P4 and P5 demonstrated higher magnitude CD8^+^ T cell responses than age-matched immunocompetent patients hospitalized with COVID-19. There are a number of possible explanations for this, the most parsimonious of which is disparate timing of sampling with respect to disease onset: d82-200 for P4 and P5 versus d13-18 for the COVID-19 controls. Thus, sampling may have occurred prior to peak T cell responses in the COVID-19 controls, or there may have been a difference in quality of the T cells elicited during acute versus late infection. Indeed, despite highly variable cellular responses to SARS-CoV-2 in COVID-19 patients, studies with larger cohorts found that a hyperactivated/exhausted T cell “immunotype” was associated with acute severe disease and that the quality of T cell responses differed in acute versus convalescent phases^35,36^. Further work is needed to characterize the phenotype of responding T cells in COVID-19 patients with B cell deficiencies.

Limitations to our study include a small number of patients and the use of convenience samples. Larger clinical studies in immunocompromised populations are needed, including serial sampling to further elucidate therapies that promote immune evasion. Our work and others’ emphasize the need to both protect immunocompromised patients from acquiring infection, and to prevent the forward spread of viruses with immune escape mutations. Such needs might be met with broad spectrum monoclonal antibodies and next generation SARS-CoV-2 vaccines that induce potent neutralizing antibody responses to prevent infection and memory CD8^+^ T cell responses to control breakthrough.

## Methods

This study was approved by the institutional review board at Emory University under protocols STUDY00000260, 00022371, and 00045821. Clinical data was obtained by electronic medical review. Residual nasopharyngeal (NP) swab and serum samples were obtained from the Emory Medical Labs. Whole blood samples were obtained after patient enrollment and consent.

### SARS-CoV-2 molecular testing, genome sequencing, and analysis

Total nucleic acid was extracted from NP swabs and tested for SARS-CoV-2 total RNA (using an N2 target) as well as subgenomic RNA as previously described^37^. Samples underwent DNase treatment (ArcticZymes, Tromso, Norway), random hexamer cDNA synthesis (Invitrogen, New England Biolabs), Nextera XT library indexing and amplification (Illumina), and Illumina sequencing. As a negative control, water was included with each batch of samples starting from DNase. As a positive control, *in vitro* transcribed ERCC spike-ins (NIST) were added to each sample prior to cDNA synthesis. In order to analyze intra-sample single nucleotide variants (iSNVs) in patients with more than one time point available, duplicate libraries were made from extracted total nucleic acid from each sample. Reads from both libraries were merged and underwent reference-based SARS-CoV-2 genome assembly using reference NC_045512.1 (viral-ngs version 2.1.19.0-rc119). Sequences from immunocompromised patients were aligned with 301 reference sequences collected from patients within the Emory Healthcare System between 1/1/2021 and 4/30/2021 using MAFFT as implemented in geneious (geneious.com). A maximum-likelihood tree was constructed using a general time reversible model with empirical base frequencies and a 3 rate model in IQ-TREE version 2.0 with 1,000 ultrafast boostraps^38^ and visualized in FigTree (http://tree.bio.ed.ac.uk/software/figtree).

To identify iSNVs, reads were mapped to reference sequence NC_045512.1 using minimap2, variants were called using vphaser2 with maximum strand bias of 5, and variants annotated with SNPeff, all as implemented in viral-ngs version 2.1.19.0-rc119. Reads containing iSNVs were manually inspected in genious (geneious.com), and iSNVs were removed from further consideration if they were primarily found in the same position across all metagenomic sequencing reads, suggesting an artefact of Nextera library construction. To minimize false-positives from PCR or sequencing error, iSNVs are only reported if they were present in two replicate libraries from each sample and had a total frequency greater than 2%^39^.

### Materials

293T cells were purchased from ATCC (CRL-3216). A HeLa cell line transduced to stably express the human ACE2 receptor (ACE2-HeLa) was generously provided by David Nemazee^17^. Anti-SARS-CoV-2 neutralizing monoclonal IgG CC12.1 was generously provided by Dennis R. Burton and the International AIDS Vaccine Initiative^17^. Purified SARS-CoV-2 cross-reactive, anti-SARS monoclonal antibody CR3022^40^ was generously provided by Jens Wrammert. Plasmids pCMV ΔR8.2 (replication defective HIV-1 backbone)^41^ pHR’ CMV-Luc (luciferase reporter plasmid)^41^ VRC7480 (expresses SARS-CoV-2 Wuhan-Hu-1 full-length spike)^42^ and TMPRSS2 (expresses human TMPRSS2)^42^ were generously provided by the Vaccine Research Center, NIAID/NIH under a Material Transfer Agreement with Emory University. Plasmid nCoV-2P-F3CH2S^43^ expressing a His-tagged, pre-fusion stabilized SARS-CoV-2 spike trimer from Wuhan-Hu-1 isolate was generously provided by Jason McLellan. Previously described peptide megapools^44^ containing 15-mers overlapping by 10 residues of the SARS-CoV-2 spike ORF (CD4-S); 15-mers overlapping by 10 residues of all other non-spike SARS-CoV-2 ORFs (CD4-R); predicted HLA class I epitopes from SARS-CoV-2, including spike; and predicted HLA class I epitopes from SARS-CoV-2, not including spike, were generously provided by Alba Grifoni, Alessandro Sette, and Daniela Weiskopf.

### SARS-CoV-2 Wuhan-Hu-1 spike trimer protein expression

Spike trimer plasmids were transiently transfected into Expi293 cells (ThermoFisher) with 5 mM kifunensine (Mfr), purified with His-Trap columns (Cytiva), trimers selected with a Superdex 200 gel filtration column (Mfr), and finished product dialyzed into 20 mM Tris pH 8.0, 200 mM sodium chloride, 0.02% sodium azide by the BioExpression and Fermentation Facility at the University of Georgia.

### Generation of autologous SARS-CoV-2 pseudovirus constructs

Q5 Site-Directed Mutagenesis Kit (New England Biolabs) was used to introduce mutations in VRC7480 corresponding to SARS-CoV-2 variant spike proteins identified in patients P1 (d30 psV), P2 (d28 psV, d39 psV, d39 + E484K psV), and P3 (d51 psV, d56 psV). Primers for site-directed mutagenesis were designed using the NEBaseChanger online tool (http://nebasechanger.neb.com/) and manufacturer recommended protocol and thermocycling conditions were followed. Incorporation of mutations was verified by Sanger sequencing. Variant open reading frames were excised from the plasmid by restriction digest and ligated separately into the parental VRC7480 plasmid using the Quick Ligation Kit (NEB). The entire spike protein open reading frame of each resulting variant plasmid was verified by Sanger sequencing, then used for large-scale DNA purification and pseudovirus production.

### PBMC isolation and use

PBMCs were separated from whole blood using BD Vacutainer® Mononuclear CPT sodium citrate tubes and stored in a liquid nitrogen tank. Cryopreserved PBMCs were thawed in a 37 °C water bath and washed with RPMI 1640 containing 10% fetal bovine serum (FBS, Gemini), 100 units/ml of penicillin, and 100 μg/ml of streptomycin (Gibco) referred to as R10 complete medium, as well as 2 units/ml RNase-free DNase I (Sigma). Before use, cells were counted and checked for viability using a Guava cell counter (Luminex).

### SARS-CoV-2 pseudovirus neutralization assay

Plasma and serum sample neutralizing activity was measured against SARS-CoV-2 pseudoviruses constructed from HIV-1 lentiviruses carrying luciferase reporter genes and pseudotyped with full-length SARS-CoV-2 spike protein. The following neutralization assay was adapted from previously published methods^42^.

#### Pseudovirus production

Pseudoviruses were produced by seeding 16 million 293T cells (ATCC CRL-3216) into DMEM with 10% heat-inactivated FBS and 1% GlutaMAX (ThermoFisher) (DMEM-10) in a T-150 flask the night prior to transfection and incubating at 37°C in a humidified 5% CO_2_ incubator. On the day of transfection, the HIV-1 lentiviral packaging plasmid, pCMV R8.2 (17.5 *µ*g); luciferase reporter plasmid, pHR’ CMV-Luc (17.5 *µ*g); VRC7480 expressing full length SARS-CoV-2 Wuhan-Hu-1 spike or patient variant spikes (1 *µ*g); and a plasmid expressing human TMPRSS2 (0.3 *µ*g) were co-transfected into cells using FuGENE6 transfection reagent (Promega). Flasks were incubated under the above conditions for 48-72 hours after transfection, and cell supernatant with pseudoviruses was removed, clarified by brief centrifugation, filtered (0.45 *µ*m), and stored in aliquots at -80°C until use.

#### Pseudovirus titration

ACE2-HeLa cells were seeded in Nunc Edge 2.0 plates (ThermoFisher) at 5,000 cells per well in DMEM-10 with 1% penicillin/streptomycin, and phosphate buffered saline (PBS) applied to the outer plate moats (replenished throughout the course of the experiment). Plates were stored at 37°C in a humidified 5% CO_2_ incubator. Approximately 24 hours later, SARS-CoV-2 pseudoviruses were diluted in a 2-fold dilution series in MEM with 5% FBS, 1% GlutaMAX, and 1% penicillin/streptomycin (MEM-5) and pre-incubated at 37°C for 45 minutes. DMEM-10 media was then removed from plates with cells and 50 *µ*l pseudovirus dilutions added onto ACE2-HeLa cells and incubated for two hours at 37°C. After the incubation, 150 *µ*l MEM-5 media was added and plates incubated an additional 72 hours at 37°C. After 72 hours, media was removed, wells washed with PBS, and 25 *µ*l Luciferase Cell Culture Lysis Reagent (Promega) added to wells with shaking for 15 minutes at room temperature. After shaking, lysates were clarified by centrifugation and 20 *µ*l lysate added to 96-well black and white isoplates (Perkin-Elmer). Fifty *µ*l luciferase substrate (Luciferase Assay System, Promega) was added and luminescence quantified in a BioTek Synergy 2.0 microplate reader using a 5 second shake and a 5 second integration time with a gain of 245 (Biotek Synergy Neo2). The mean background signal (average of signal in cell only wells) was subtracted from the signal of wells with pseudoviruses prior to determining mean pseudovirus relative light units (RLUs). The final dilution of SARS-CoV-2 pseudoviruses yielding approximately 1,000,000-2,000,000 RLUs was selected for future experiments.

#### Neutralization assay

The same protocol was followed as for the titration above, except that pseudoviruses at 2X the final dilution were mixed with equal parts sample in a dilution series (1:15 starting dilution to achieve a final starting dilution of 1:30) in triplicate and preincubated at 37°C for 45 minutes in a humidified 5% CO_2_ incubator. To compare the ability of a serum, plasma, or mAb to neutralize different psV, the same input RLUs were used for each psV (e.g., 1,000,000 RLU of P1 d30 psV and 1,000,000 RLUs of Wuhan-Hu-1 psV).The pseudovirus-sample mixture was plated onto ACE2-HeLa cells and incubated for 2 hours at 37°C. Mean background signal (cell only wells) was subtracted from signal of wells containing pseudovirus or pseudovirus plus sample. Percent neutralization was determined using the following formula:

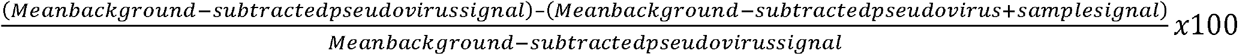

Percent neutralization curves were fitted with a 3-parameter non-linear regression (Prism v8) to determine the half-maximal inhibitory concentration (IC50).

Samples that did not achieve 50% neutralization at 1:30 were assigned a reciprocal titer of ≤10 and were confirmed in triplicate at a single sample dilution of 1:30 in a separate, independent experiment (except that there was only enough P2 d55 serum to test at a single 1:60 dilution in a second experiment). Samples with >50% neutralization at 1:30 were analyzed in two or more independent experiments with full serial dilutions. The SARS-CoV-2 neutralizing monoclonal antibody CC12.1 was used as a positive control for each experiment. For the negative control, sera pooled from six healthy subjects was utilized, where the individual negative control sera were collected in Atlanta, Georgia in March to April 2020 from persons with no COVID-19 history.

### SARS-CoV-2 Spike Trimer Capture ELISA

The following ELISA was adapted from previously published methods^17^: 96-well half area, high binding plates (Corning #3690) were coated with anti-6x-His-tag monoclonal antibody (#MA1-21315MG, ThermoFisher) at 2 *µ*g /mL in PBS at 4°C overnight. After washing three times in PBS with 0.05% Tween (wash buffer), plates were blocked with 3% BSA in PBS for 1 hour at room temperature (RT). His-tagged spike trimers at 5 *µ*g /mL in PBS with 1% BSA and 0.05% Tween (diluent) were incubated on plates for 90 minutes at RT. Plates were washed before heat-inactivated subject serum/plasma sample dilutions were applied to the plates for 90 minutes at RT. CR3022 was applied as a positive control. Pooled control sera from the six seronegative donors described above served as the negative control. After sample incubation, plates were washed and alkaline phosphatase-conjugated goat-anti-human IgG (#109-055-008, Jackson ImmunoResearch) in diluent was applied for 1 hour at RT. After washing, plates were developed with phosphatase substrate (Sigma) in staining buffer (40 mM sodium carbonate and 10 mM magnesium chloride hexahydrate, pH 9.8). Absorbance was measured in a BioTek Synergy 2.0 microplate reader at 405 nm. Data were background-subtracted with absorbance from blank wells. Healthy control cutoffs were determined by measuring absorbances from six healthy control subject samples (described above) and applying the formula: Cutoff = mean + (standard deviation * 2.177)^45^.

Mean background-subtracted absorbances were plotted relative to sample dilutions and curves fitted with a four-parameter non-linear regression (Prism v8). To ascertain a precise endpoint titer (ET), curve data (best fit values for the bottom, top, logEC50, and hill slope) were processed by a MATLAB program designed to determine the sample dilution at which each regression curve intersected the healthy control cutoff value. Samples with background-subtracted absorbances below the healthy control cutoff were assigned an ET of 10. Samples with background-subtracted absorbances slightly above the healthy control cutoff but with poor curve fitting due to low signal were assigned an ET of 30. All samples were analyzed in at least two independent experiments.

### Immunophenotyping by flow cytometry

Thawed Cryopreserved PBMCs were directly used for phenotypic staining. Approximately one million viable PBMCs were stained with Zombie aqua fixable cell viability dye (BioLegend) to exclude dead cells; washed with PBS containing 2% FBS, referred to as FACS buffer; surface-stained with the following fluorescent monoclonal antibodies: CD3 (clone SK7, BioLegend), CD4 (clone SK3, BioLegend), CD8 (clone SK1, BioLegend), CD19 (clone HIB19, eBioscience), CD20 (clone 2H7, BioLegend), CD45RA (clone HI100, BD), CCR7 (clone G043H7, BioLegend), CD27 (clone M-T271, BioLegend), CD38 (clone HB7, BioLegend). Flow cytometry data were collected on an LSR Fortessa (BD Biosciences) and analyzed using FlowJo software V10 (Tree Star). Patient P2 only had 0.5 million PBMC available for staining.

### Intracellular Cytokine Staining

For measuring SARS-CoV-2-specific CD4 and CD8 T cell responses, thawed cryopreserved PBMCs were rested overnight in a 5% CO_2_ incubator at 37°C in R10 with DNase I. On day 2, approximately 1-2 million viable PBMCs per sample were stimulated for two hours at 37 °C in R10 with 1 *µ*g/ml of CD4-S, CD4-R, CD8-A, or CD8-B, or negative control (R10 with equivalent peptide vehicle (DMSO)), or positive control (R10 with 1 *µ*g/ml Staphylococcal enterotoxin B (SEB, Sigma)) in the presence of anti-CD28 and anti-CD49d (BD Biosciences). After two hours, a cocktail protein transport inhibitor (eBioscience) was added and cells were cultured for an additional 4 hours at 37°C, then stored at 4°C overnight. On day 3, samples were stained with aqua cell viability dye to exclude dead cells, surface stained with CD3 (clone SK7, BioLegend), CD4 (clone SK3, BioLegend), CD8 (clone SK1, BioLegend), CCR7 (clone G043H7, BioLegend), and CD45RA (clone HI100, BD) for 25 minutes. After washing with FACS buffer and fixing and permeabilizing cells with Cytofix/Cytoperm (BD Biosciences), the cells were stained intracellularly with the following fluorescent monoclonal antibodies: CD154 (clone CD40L 24-31, BioLegend), IL-2 (clone MQ1-17H12, BD Biosciences), IFN-γ (clone 4S.B3, eBioscience), TNF (clone Mab11, BD Biosciences). Flow cytometry data were collected on an LSR Fortessa (BD Biosciences) and analyzed using FlowJo software V10 (Tree Star).

### Interferon gamma ELISPOT

Interferon gamma (IFN-γ) ELISPOT was used to enumerate the number of individual T cells secreting IFN-γ after approximately 0.2 million thawed cryopreserved PBMCs were stimulated with antigen. 96-well ELISPOT filter plates (Millipore, #MSIPS4W10) were coated with anti-human-IFN-γ (clone 1-D1K, Mabtech) overnight at 4°C. Plates were washed and blocked with R10 1-2 hours at 37°C in a 5% CO_2_ incubator prior to use. Thawed and rested PBMC were resuspended in R10 at 0.2 million cells/well and mixed with 1 μg/ml each of both SARS-CoV-2 CD4-S and CD4-R peptide megapools, a negative control (R10 only), or positive control (1 μg/ml SEB) in the presence of anti-CD28 and anti-CD49d and distributed into ELISPOT plates and incubated 21-24 hours at 37 °C. IFN-γ spots were detected with biotinylated murine anti-human IFN-γ antibody (clone 7-B6-1, Mabtech), followed by incubation with streptavidin-HRP (BD) and then developed using AEC substrate (EMD Millipore). Developed and dried ELISPOT plates were scanned and counted by using an automated ELISPOT counter (Cellular Technologies Limited). Each spot forming unit (SFU) indicates an IFN-γ secreting cell and is reported as the number of SFU per million PBMC.

## Data Availability

All sequence data (cleaned of human reads) are available on NCBI under BioProject PRJNA634356. SARS-CoV-2 consensus sequences are available in GISAID under accession numbers EPI_ISL_1503958 and EPI_ISL_6913932-43.

## Funding

This study was supported by CDC contract 75D30121C10084 under BAA ERR 20-15-2997 (AB, JW, KK, AP), National Institutes of Health (NIH) grant 5UM1AI148576-02 (NR and EMS), the Pediatric Research Alliance Center for Childhood Infections and Vaccines and Children’s Healthcare of Atlanta, and the Emory Woodruff Health Sciences Center COVID-19 Urgent Research Engagement (CURE) Center, made possible by generous philanthropic support from the O. Wayne Rollins Foundation and the William Randolph Hearst Foundation. Research reported in this publication was supported by the National Institute of Allergy and Infectious Diseases of the NIH under Award Number K08AI139348 (AP) and NIH contract No. 75N9301900065 (DW). The content is solely the responsibility of the authors and does not necessarily represent the official views of the NIH. LJI has filed for patent protection for various aspects of T cell epitope and vaccine design work.

## Acknowledgements

We thank the patients and families who participated in this study, as well as all clinical staff who participated in the care of these patients. We also thank members of the Emory Clinical Virology Laboratory and Georgia Clinical Research Centers for assistance with sample collection and processing, members of the University of Georgia’s BioExpression and Fermentation Facility for spike protein expression and purification, and Dennis Burton, Alba Grifoni, Jason McLellan, David Nemazee, Alessandro Sette, Jens Wrammert, and the NIH Vaccine Research Center for generously sharing reagents. This study was supported in part by the Emory Integrated Genomics Core (EIGC) and Emory Integrated Computational Core (EICC), which are subsidized by the Emory University School of Medicine and is one of the Emory Integrated Core Facilities.

## Figures

**Fig. S4:**
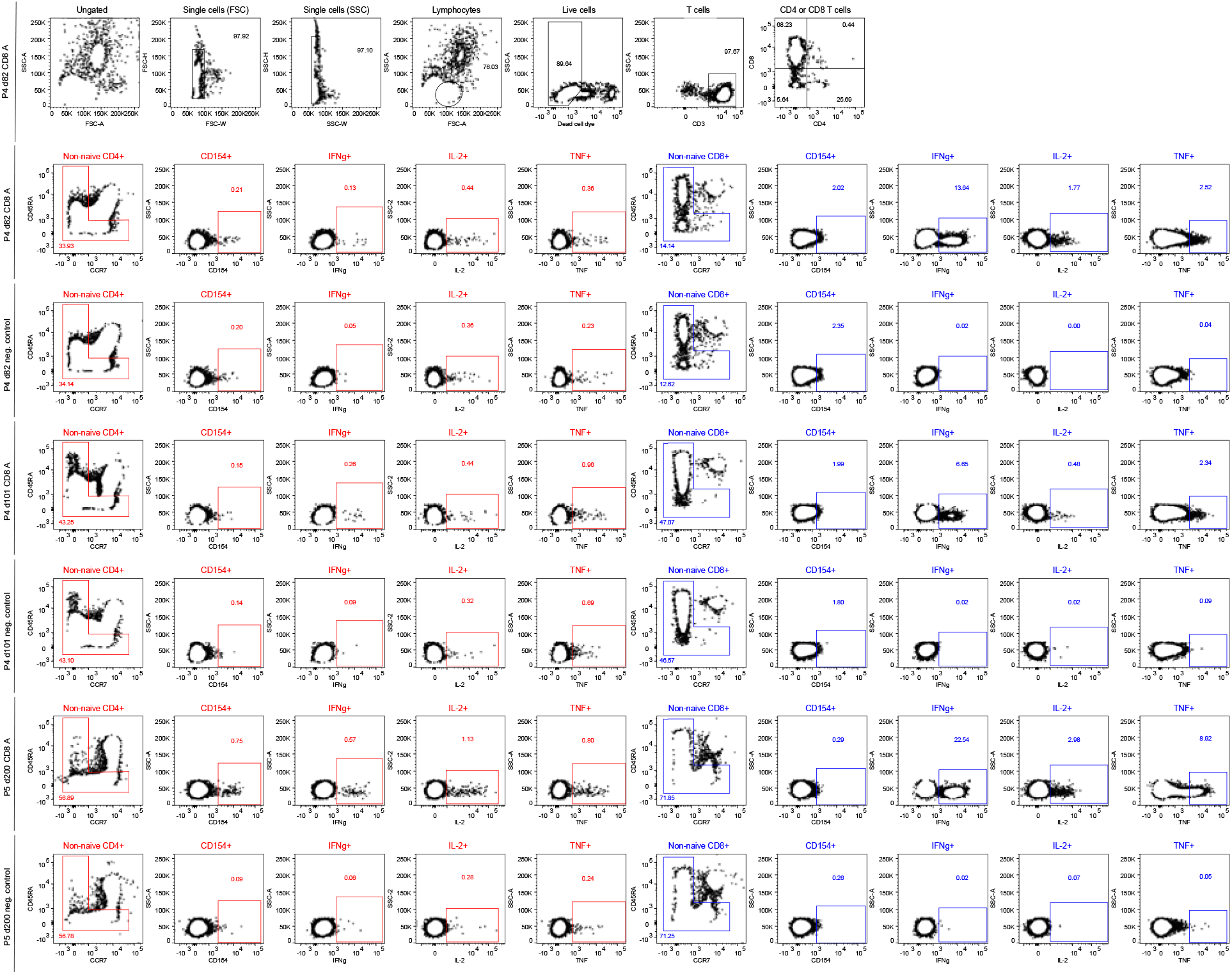
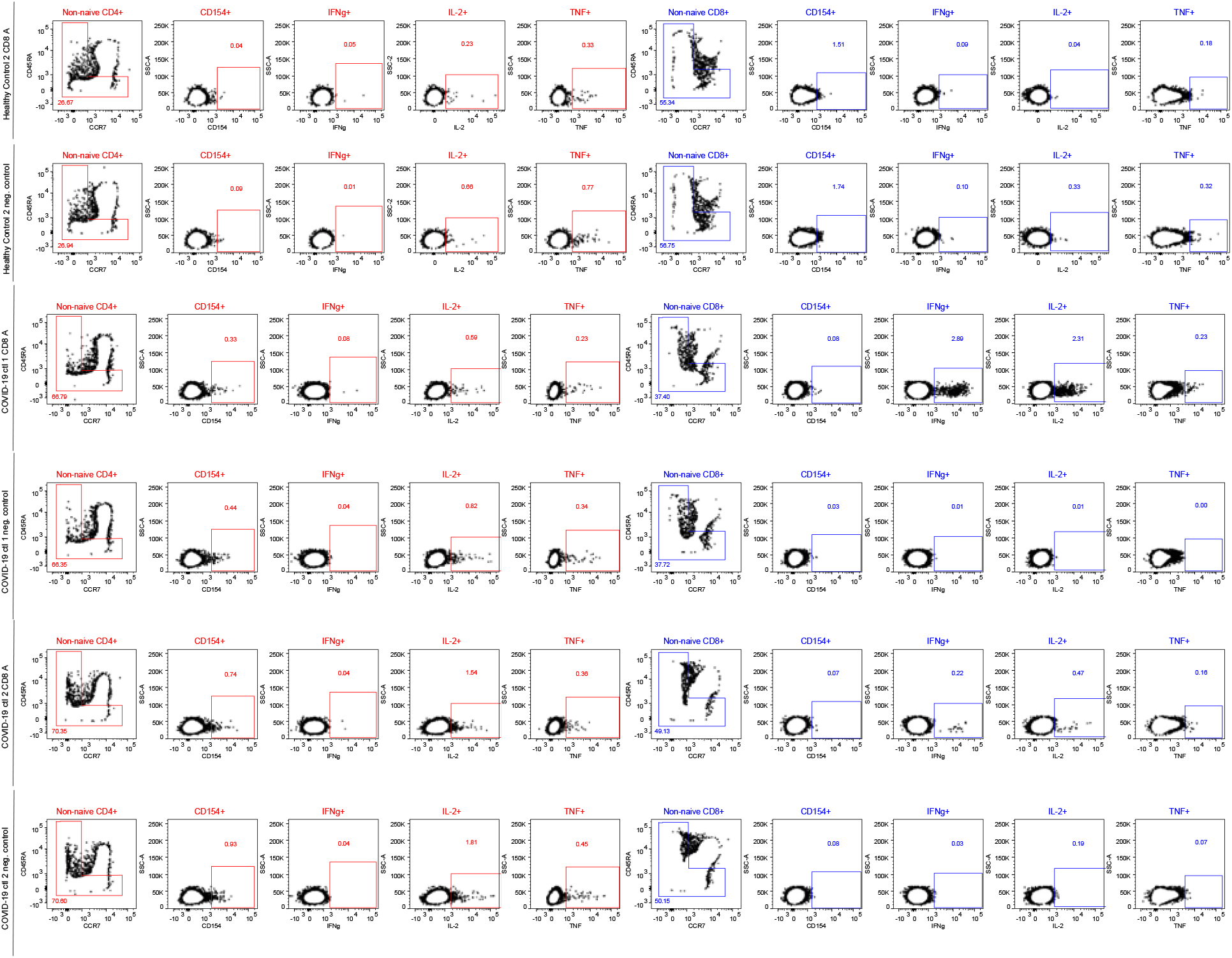
Gating strategy used to identify SARS-CoV-2-specific CD4^+^ and CD8^+^ T cells. Frequencies of non-naive CD4^+^ or CD8^+^ T cells responding to stimulation with a negative control (media with equivalent peptide vehicle) or peptide megapool containing predicted CD8+ T cell epitopes of SARS-CoV-2 ORFs (including spike) as an example of antigen-specific responses, were identified by intracellular cytokine staining and flow cytometry.

## Notes

### Competing Interest Statement

The authors have declared no competing interest.

